# Viral dynamics of acute SARS-CoV-2 infection

**DOI:** 10.1101/2020.10.21.20217042

**Authors:** Stephen M. Kissler, Joseph R. Fauver, Christina Mack, Scott W. Olesen, Caroline Tai, Kristin Y. Shiue, Chaney C. Kalinich, Sarah Jednak, Isabel M. Ott, Chantal B.F. Vogels, Jay Wohlgemuth, James Weisberger, John DiFiori, Deverick J. Anderson, Jimmie Mancell, David D. Ho, Nathan D. Grubaugh, Yonatan H. Grad

## Abstract

**Background:** SARS-CoV-2 infections are characterized by viral proliferation and clearance phases and can be followed by low-level persistent viral RNA shedding. The dynamics of viral RNA concentration, particularly in the early stages of infection, can inform clinical measures and interventions such as test-based screening.

**Methods:** We used prospective longitudinal RT-qPCR testing to measure the viral RNA trajectories for 68 individuals during the resumption of the 2019-20 National Basketball Association season. For 46 individuals with acute infections, we inferred the peak viral concentration and the duration of the viral proliferation and clearance phases.

**Findings:** According to our mathematical model, we found that viral RNA concentrations peaked an average of 3.3 days (95% credible interval [2.5, 4.2]) after first possible detectability at a cycle threshold value of 22.3 [20.5, 23.9]. The viral clearance phase lasted longer for symptomatic individuals (10.9 days [7.9, 14.4]) than for asymptomatic individuals (7.8 days [6.1, 9.7]). A second test within 2 days after an initial positive PCR substantially improves certainty about a patient’s infection phase. The effective sensitivity of a test intended to identify infectious individuals declines substantially with test turnaround time.

**Conclusions:** SARS-CoV-2 viral concentrations peak rapidly regardless of symptoms. Sequential tests can help reveal a patient’s progress through infection stages. Frequent rapid-turnaround testing is needed to effectively screen individuals before they become infectious.

## Introduction

A critical strategy to curb the spread of SARS-CoV-2 is to rapidly identify and isolate infectious individuals. Because symptoms are an unreliable indicator of infectiousness and infections are frequently asymptomatic^1^, testing is key to determining whether a person is infected and may be contagious. Real time quantitative reverse transcriptase polymerase chain reaction (RT-qPCR) tests are the gold standard for detecting SARS-CoV-2 infection. Normally, these tests yield a binary positive/negative diagnosis based on detection of viral RNA. However, they can also quantify the viral titer via the cycle threshold (Ct). The Ct is the number of thermal cycles needed to amplify sampled viral RNA to a detectable level: the higher the sampled viral RNA concentration, the lower the Ct. This inverse correlation between Ct and viral concentration makes RT-qPCR tests far more valuable than a binary diagnostic, as they can be used to reveal a person’s progress through key stages of infection^2^, with the potential to assist clinical and public health decision-making. However, the dynamics of the Ct during the earliest stages of infection, when contagiousness is rapidly increasing, have been unclear, because diagnostic testing is usually performed after the onset of symptoms, when viral RNA concentration has peaked and already begun to decline, and performed only once^3,4^. Without a clear picture of the course of SARS-CoV-2 viral concentrations across the full duration of acute infection, it has been impossible to specify key elements of testing algorithms such as the frequency of rapid at-home testing^5^ that will be needed to reliably screen infectious individuals before they transmit infection. Here, we fill this gap by analyzing the prospective longitudinal SARS-CoV-2 RT-qPCR testing performed for players, staff, and vendors during the resumption of the 2019-20 National Basketball Association (NBA) season.

## Methods

### Data collection

The study period began in teams’ local cities from June 23^rd^ through July 9^th^, 2020, and testing continued for all teams as they transitioned to Orlando, Florida through September 7^th^, 2020. A total of 68 individuals (90% male) were tested at least five times during the study period and recorded at least one positive test with Ct value <40. Most (85%) of consecutive tests were recorded within one day of each other and fewer than 3% of the intervals between consecutive tests exceeded 4 days (**Supplemental Figure 1**). Many individuals were being tested daily as part of Orlando campus monitoring. Due to a lack of new infections among players and team staff after clearing quarantine in Orlando, all players and team staff included in the results pre-date the Orlando phase of the restart. A diagnosis of “acute” or “persistent” infection was abstracted from physician records. “Acute” denoted a likely new infection. “Persistent” indicated the presence of virus in a clinically recovered individual, likely due to infection that developed prior to the onset of the study. There were 46 acute infections; the remaining 22 individuals were assumed to be persistently shedding SARS-CoV-2 RNA due to a known infection that occurred prior to the study period^6^. This persistent RNA shedding can last for weeks after an acute infection and likely represented non-infectious viral RNA^7^. Of the individuals included in the study, 27 of the 46 with acute infections and 40 of the 68 overall were from staff and vendors. The Ct values for all tests for the 68 individuals included in the analysis with their designations of acute or persistent infection are depicted in **Supplemental Figures 2–5**. A schematic diagram of the data collection and analysis pipeline is given in **Figure 1**.

**Figure 1.**
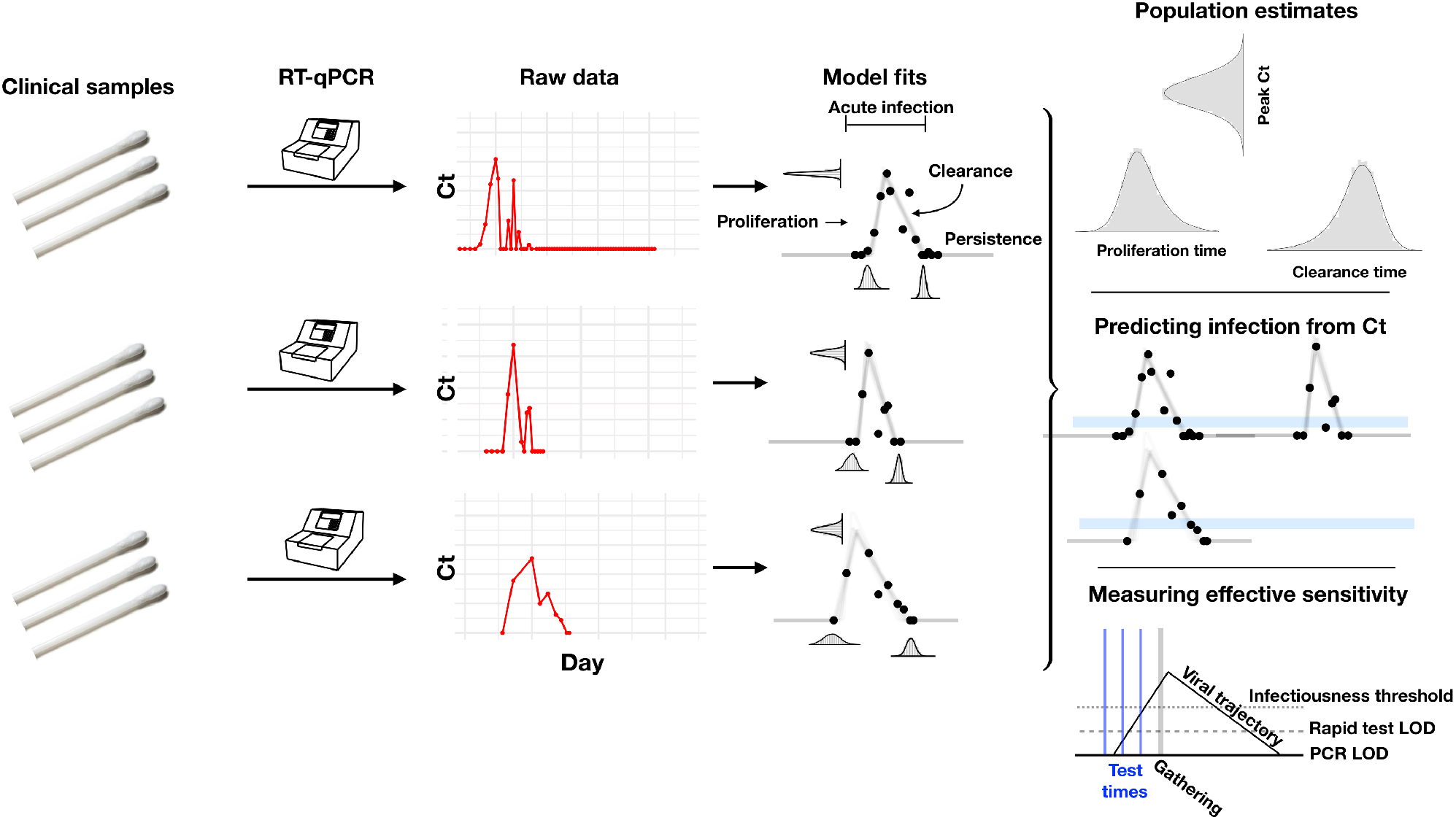
Illustration of the analysis pipeline. Combined anterior nares and oropharyngeal swabs were tested using a RT-qPCR assay to generate longitudinal Ct values (‘Raw data’, red points) for each person. Using a statistical model (see **Supplemental Figure 6** for a schematic of the model), we estimated Ct trajectories consistent with the data, represented by the thin lines under the ***‘Model fits’*** heading. These produced posterior probability distributions for the peak Ct value, the duration of the proliferation phase (first potential detectability of infection to peak Ct), and the duration of the clearance phase (peak Ct to resolution of acute infection) for each person. We estimated population means for these quantities (under heading “Population estimates”). The model fits also allowed us to determine how frequently a given Ct value or pair of Ct values within a five-unit window (blue bars, under heading “Predicting infection from Ct”) was associated with the proliferation phase, the clearance phase, or a persistent infection. Finally, the model fits allowed us to measure the ‘effective sensitivity’ of a test for predicting future infectiousness. The schematic illustration titled ***‘Measuring effective sensitivity’*** depicts the relationship between testing lags and the ability to detect infectious individuals at a gathering. The illustrated viral trajectory surpasses the infectiousness threshold (dotted line) at the time of the gathering (vertical grey bar), so unless this individual is screened by a pre-gathering test, s/he would attend the event while infectious. One day prior to the gathering, the individual could be detected by either a rapid test or a PCR test. Two days prior to the event, the individual could be detected by a PCR test but not by a rapid test. Three days prior to the event, neither test would detect the individual.

**Figure 2.**
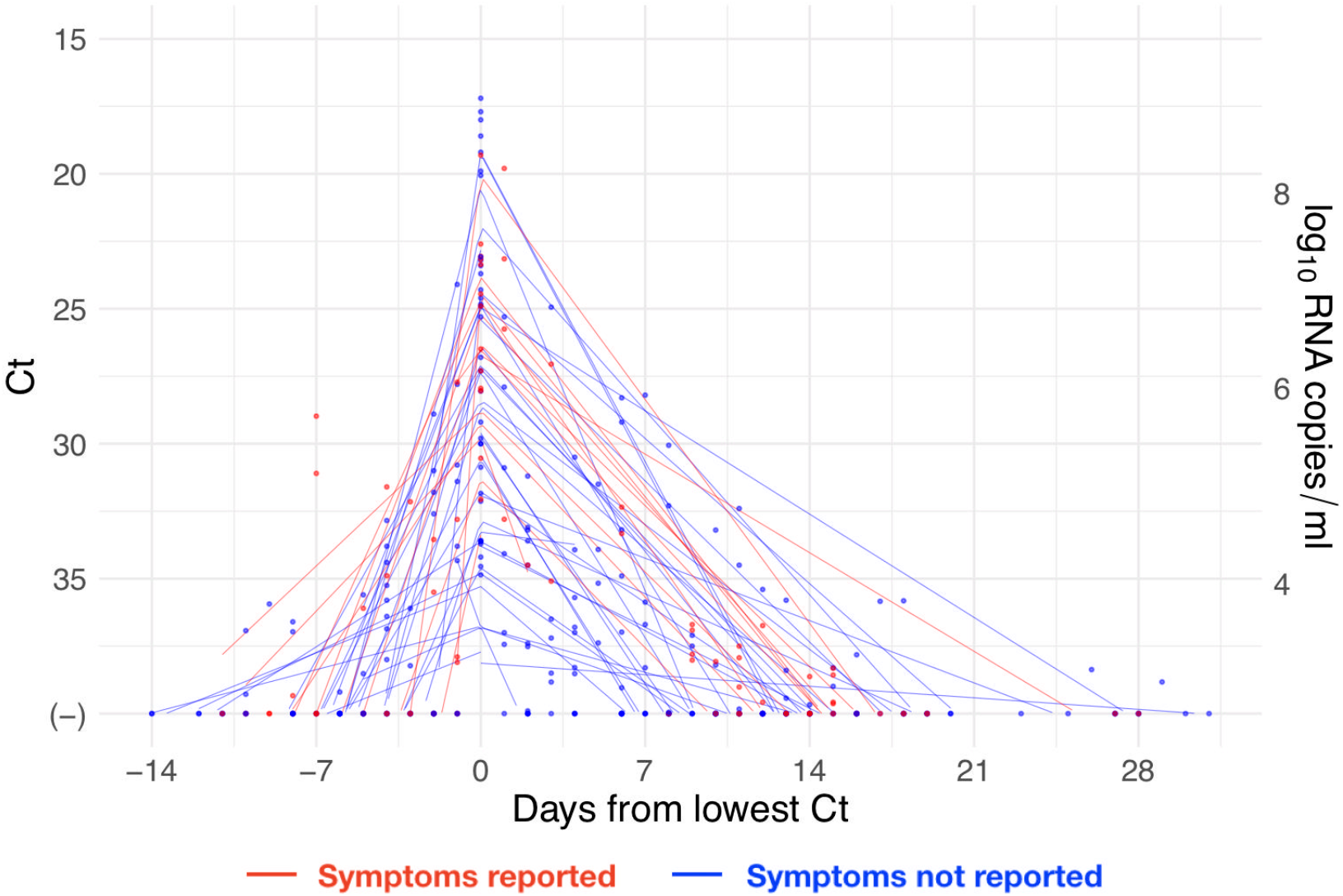
Reported Ct values with individual-level piecewise linear fits. Ct values (points) for the 46 acute infections aligned by the date when the minimum Ct was recorded for each individual. Lines depict the best-fit piecewise linear regression lines for each individual with breakpoint at day 0. Red points/lines represent individuals who reported symptoms and blue points/lines represent individuals who did not report symptoms. Five positive tests were omitted that occurred >20 days prior to the individual’s minimum Ct value, all of which had Ct > 35. The vertical axis on the right-hand side gives the conversion from Ct values to RNA concentration.

### Statistical analysis

Due to imperfect sampling, persistent viral shedding, and test uncertainty near the limit of detection, a straightforward analysis of the data would be insufficient to reveal the duration and peak magnitude of the viral trajectory. Imperfect sampling would bias estimates of the peak viral concentration towards lower concentrations/higher Ct values since the moment of peak viral concentration is unlikely to be captured. Persistent shedding and test uncertainty would bias estimates of the trajectory duration towards longer durations of infection. To address these problems, we used a Bayesian statistical model to infer the peak Ct value and the durations of the proliferation and clearance stages for the 46 acute infections (**Figure 1**; **Supplemental Methods**). We assumed that the viral concentration trajectories consisted of a proliferation phase, with exponential growth in viral RNA concentration, followed by a clearance phase characterized by exponential decay in viral RNA concentration^8^. Since Ct values are roughly proportional to the negative logarithm of viral concentration^2^, this corresponds to a linear decrease in Ct followed by a linear increase. We therefore constructed a piecewise-linear regression model to estimate the peak Ct value, the time from infection onset to peak (*i*.*e*., the duration of the proliferation stage), and the time from peak to infection resolution (*i*.*e*., the duration of the clearance stage). This allowed us to separate the viral trajectories into the three distinct phases of proliferation (from the onset of detectability to the peak viral concentration, or t_o_ to t_p_ in **Supplemental Figure 6**), clearance (from the peak viral concentration to the resolution of acute infection, or t_p_ to t_r_ in **Supplemental Figure 6**), and persistence (lasting indefinitely after the resolution of acute infection, or after t_r_ in **Supplemental Figure 6**; see also **Figure 1**). Note that for the 46 individuals with acute infections, the persistence phase is identified using the viral trajectory model, whereas for the 22 other infections the entire series of observations was classified as ‘persistent’ due to clinical evidence of a probable infection prior to the start of the study period. We estimated the parameters of the regression model by fitting to the available data using a Hamiltonian Monte Carlo algorithm^9^ yielding simulated draws from the Bayesian posterior distribution for each parameter. Full details on the fitting procedure are given in the **Supplemental Methods**. Code is available at https://github.com/gradlab/CtTrajectories.

### Inferring stage of infection

Next, we determined whether individual or paired Ct values can reveal whether an individual is in the proliferation, clearance, or persistent stage of infection. To assess the predictive value of a single Ct value, we extracted all observed Ct values within a 5-unit window (*e*.*g*., between 30 and 35 Ct) and measured how frequently these values sat within the proliferation stage, the clearance stage, or the persistent stage. We measured these frequencies across 10,000 posterior parameter draws to account for that fact that Ct values near stage transitions (*e*.*g*., near the end of the clearance stage) could be assigned to different infection stages depending on the parameter values (see **Figure 1**, bottom-right). We did this for 23 windows with midpoint spanning from Ct = 37.5 to Ct = 15.5 in increments of 1 Ct.

To calculate the probability that a Ct value sitting within the 5-unit window corresponded to an acute infection (*i*.*e*., either the proliferation or the clearance stage), we summed the proliferation and clearance frequencies for all samples within that window and divided by the total number of samples in the window. We similarly calculated the probability that a Ct sitting within the 5-unit window corresponded to just the proliferation phase.

To assess the information gained by conducting a second test within two days of an initial positive, we restricted our attention to all samples that had a subsequent sample taken within two days. We repeated the above calculations for (a) consecutive tests with decreasing Ct and (b) consecutive tests with increasing Ct. That is, we measured the frequency with which a given Ct value sitting within a 5-unit window, followed by a second test with either lower or higher Ct, sat within with the proliferation, clearance, or persistence stages.

### Measuring the effective sensitivity of screening tests

The sensitivity of a test is defined as the probability that the test correctly identifies an individual who is positive for some criterion of interest. For clinical diagnostic SARS-CoV-2 tests, the criterion of interest is current infection with SARS-CoV-2. Alternatively, a common goal is to predict infectiousness at some point in the future, as in the context of test-based screening prior to a social gathering. The ‘effective sensitivity’ of a test in this context (*i*.*e*., its ability to predict future infectiousness) may differ substantially from its clinical sensitivity (*i*.*e*., its ability to detect current infection). A test’s effective sensitivity depends on its inherent characteristics, such as its limit of detection and sampling error rate, as well as the viral dynamics of infected individuals.

To illustrate this, we estimated the effective sensitivity of (a) a test with limit of detection of 40 Ct and a 1% sampling error probability (akin to RT-qPCR), and (b) a test with limit of detection of 35 Ct and a 5% sampling error probability (akin to some rapid antigen tests). We measured the frequency with which such tests would successfully screen an individual who would be infectious at the time of a gathering when the test was administered between 0 and 3 days prior to the gathering, given viral trajectories informed by the longitudinal testing data (see schematic in **Figure 1**). To accomplish this, we drew 1,000 individual-level viral concentration trajectories from the fitted model, restricting to trajectories with peak viral concentration above a given infectiousness threshold (any samples with peak viral concentration below the infectiousness threshold would never be infectious and so would not factor into the sensitivity calculation). For the main analysis, we assumed that the infectiousness threshold was at 30 Ct^10^. In a supplementary analysis, we also assessed infectiousness thresholds of 35 and 20 Ct. We drew onset-of-detectability times (*i*.*e*., the onset of the proliferation stage) according to a random uniform distribution so that each person would have a Ct value exceeding the infectiousness threshold at the time of the gathering. Then, we calculated the fraction of trajectories that would be successfully screened using (a) a test with limit of detection of 40 Ct and (b) a limit of detection of 35 Ct, administered between 0 and 3 days prior to the gathering. Full details are given in the **Supplemental Methods** and **Supplemental Figure 7A**.

Next, we shifted attention from the individual to the gathering. We estimated the number of individuals who would be expected to arrive at a 1,000-person gathering while infectious given each testing strategy (40 Ct limit of detection with 1% false negative rate; 35 Ct limit of detection with 5% false negative rate) assuming a 2% prevalence of PCR-detectable infection in the population. To do so, we again drew 1,000 individual-level viral concentration trajectories from the fitted model and drew onset-of-detectability times according to a random uniform distribution from the range of possible times that would allow for the person to have detectable virus (Ct < 40) during the gathering. We counted the number of people who would have been infectious at the gathering (a) in the absence of testing and (b) given a test administered between 0 and 3 days prior to the gathering. As before, we assumed that infectiousness corresponded to a Ct value of 30 for the main analysis and considered infectiousness thresholds of 35 Ct and 20 Ct in a supplemental analysis. Full details are given in the **Supplemental Methods** and **Supplemental Figure 7B**. To facilitate the exploration of different scenarios, we have generated an online tool (https://stephenkissler.shinyapps.io/shiny/) where users can input test and population characteristics and calculate the effective sensitivity and expected number of infectious individuals at a gathering.

## Results

Of the 46 individuals with acute infections, 13 reported symptoms at the time of diagnosis; the timing of the onset of symptoms was not recorded. The median number of positive tests for the 46 individuals was 3 (IQR [2, 5]). The minimum recorded Ct value across the 46 individuals had mean 26.4 (IQR [23.2, 30.4]). The recorded Ct values for the acute infections with individual-level piecewise linear regressions are depicted in **Figure 2**.

Based on the viral trajectory model, the mean peak Ct value for symptomatic individuals was 22.3 (95% credible interval [19.3, 25.3]), the mean duration of the proliferation phase was 3.4 days [2.0, 4.8], and the mean duration of clearance was 10.9 days [7.9, 14.4] (**Figure 3**). This compares with 22.3 Ct [20.0, 24.4], 3.5 days [2.5, 4.5], and 7.8 days [6.1, 9.7], respectively, for individuals who did not report symptoms at the time of diagnosis (**Figure 3**). This yielded a slightly longer overall duration of acute infection for individuals who reported symptoms (14.3 days [11.0, 17.7]) *vs*. those who did not (11.2 days [9.4, 13.4]). For all individuals regardless of symptoms, the mean peak Ct value, proliferation duration, clearance duration, and duration of acute shedding were 22.3 Ct [20.5, 23.9], 3.3 days [2.5, 4.2], 8.5 days [6.9, 10.1], and 11.7 days [10.1, 13.6] (**Supplemental Figure 8**). A full list of the model-inferred viral trajectory parameters is reported in **Table 1**. There was a substantial amount of individual-level variation in the peak Ct value and the proliferation and clearance stage durations (**Supplemental Figures 9–14**).

**Table 1.**
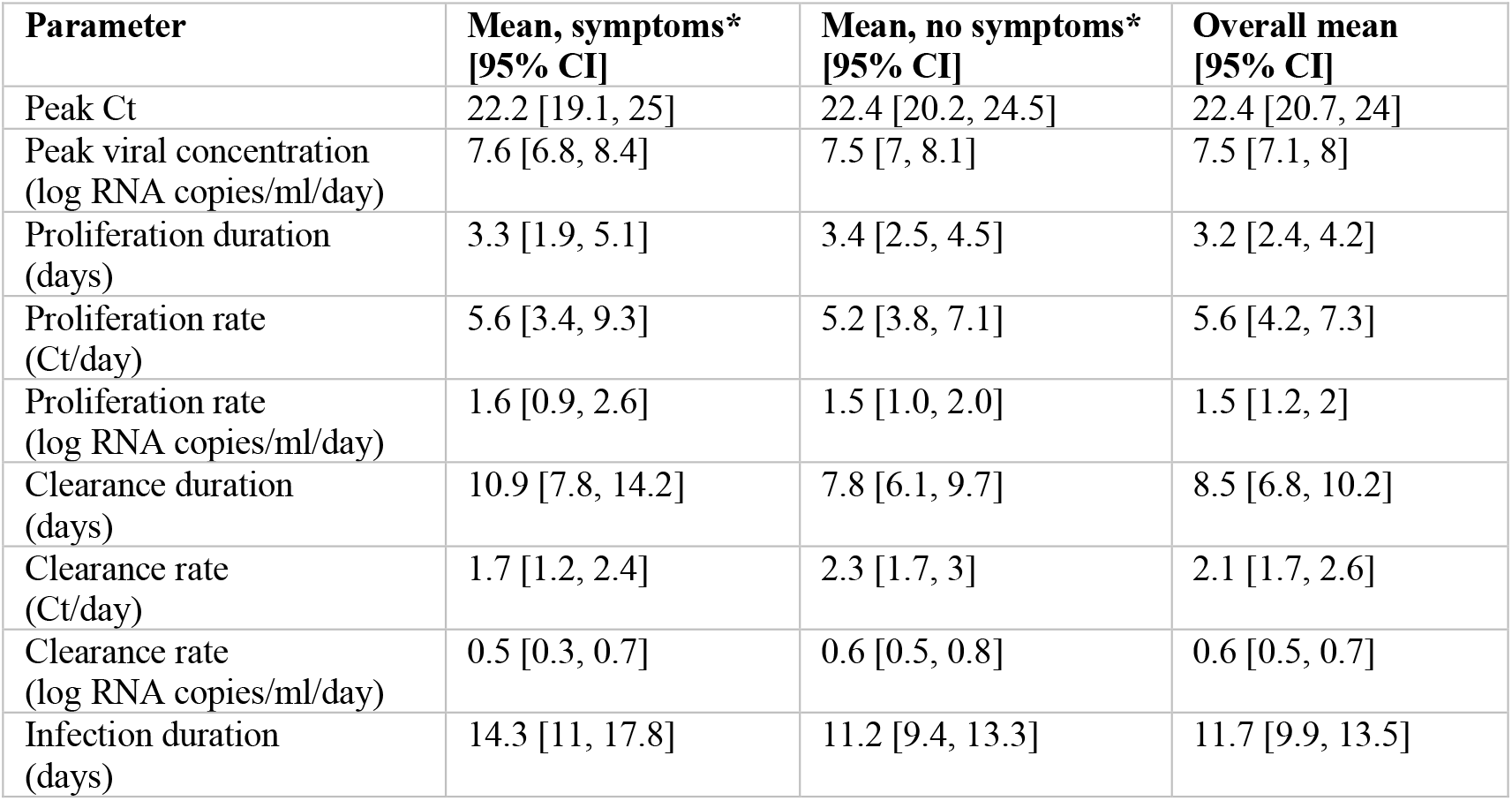
Viral dynamic parameters, overall and separated by reported symptoms. Population sizes for each category are: symptoms, *N=*13; no symptoms, *N=*33; overall, *N=*46. **\***Symptom reporting was imperfect as follow-up during course of disease was not systematic for all individuals.

| Parameter | Mean, symptoms*<br>[95% CI] | Mean, no symptoms*<br>[95% CI] | Overall mean<br>[95% CI] |
| --- | --- | --- | --- |
| Peak Ct | 22.2 [19.1, 25] | 22.4 [20.2, 24.5] | 22.4 [20.7, 24] |
| Peak viral concentration<br>(log RNA copies/ml/day) | 7.6 [6.8, 8.4] | 7.5 [7, 8.1] | 7.5 [7.1, 8] |
| Proliferation duration<br>(days) | 3.3 [1.9, 5.1] | 3.4 [2.5, 4.5] | 3.2 [2.4, 4.2] |
| Proliferation rate<br>(Ct/day) | 5.6 [3.4, 9.3] | 5.2 [3.8, 7.1] | 5.6 [4.2, 7.3] |
| Proliferation rate<br>(log RNA copies/ml/day) | 1.6 [0.9, 2.6] | 1.5 [1.0, 2.0] | 1.5 [1.2, 2] |
| Clearance duration<br>(days) | 10.9 [7.8, 14.2] | 7.8 [6.1, 9.7] | 8.5 [6.8, 10.2] |
| Clearance rate<br>(Ct/day) | 1.7 [1.2, 2.4] | 2.3 [1.7, 3] | 2.1 [1.7, 2.6] |
| Clearance rate<br>(log RNA copies/ml/day) | 0.5 [0.3, 0.7] | 0.6 [0.5, 0.8] | 0.6 [0.5, 0.7] |
| Infection duration<br>(days) | 14.3 [11, 17.8] | 11.2 [9.4, 13.3] | 11.7 [9.9, 13.5] |

**Figure 3.**
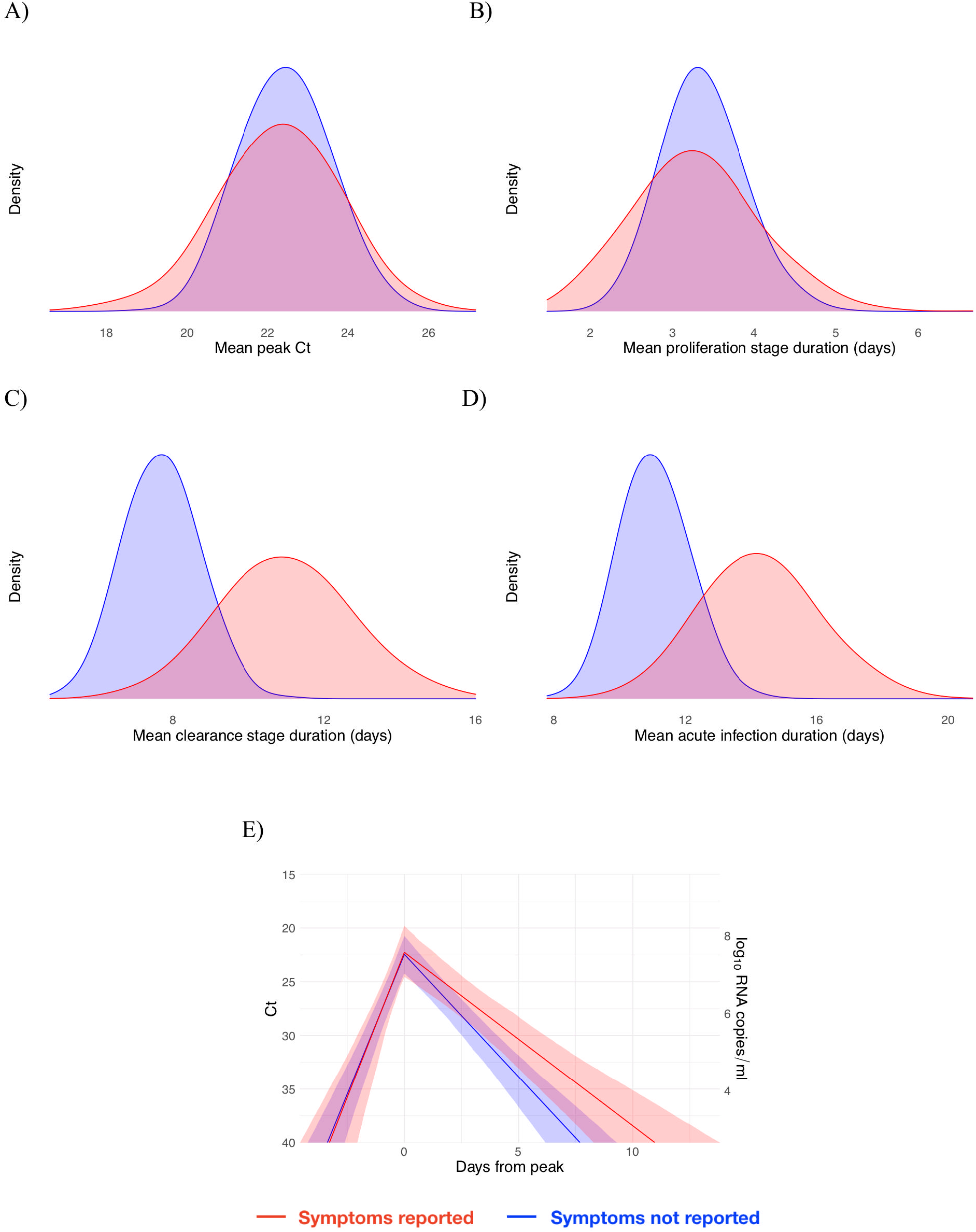
Peak Ct value and infection stage duration distributions according to symptoms reported at time of diagnosis. Posterior distributions obtained from 2,000 simulated draws from the posterior distributions for mean peak Ct value (A), mean duration of the proliferation stage (first potential infection detectability to peak Ct, B), mean duration of the clearance stage (peak Ct to resolution of acute RNA shedding, C), and total duration of acute shedding (D) across the 46 individuals with an acute infection. The distributions are separated according to whether the person reported symptoms (red, 13 individuals) or did not report symptoms (blue, 33 individuals). The mean Ct trajectory corresponding to the mean values for peak Ct, proliferation duration, and clearance duration for symptomatic *vs*. asymptomatic individuals is depicted in (E) (solid lines), where shading depicts the 90% credible intervals.

Using the full dataset of 68 individuals, we estimated the frequency with which a given Ct value was associated with an acute infection (*i*.*e*., the proliferation or clearance phase, but not the persistence phase), and if so, the probability that it was associated with the proliferation stage alone. The probability of an acute infection increased rapidly with decreasing Ct (increasing viral load), with Ct < 30 virtually guaranteeing an acute infection in this dataset (**Figure 4A**). However, a single Ct value provided little information about whether an acute infection was in the proliferation or the clearance stage (**Figure 4B**). This is unsurprising since the viral trajectory must pass through any given value during both the proliferation and the clearance stage. With roughly uniform sampling over time, a given Ct value is more likely to correspond to the clearance stage simply because the clearance stage is longer.

**Figure 4.**
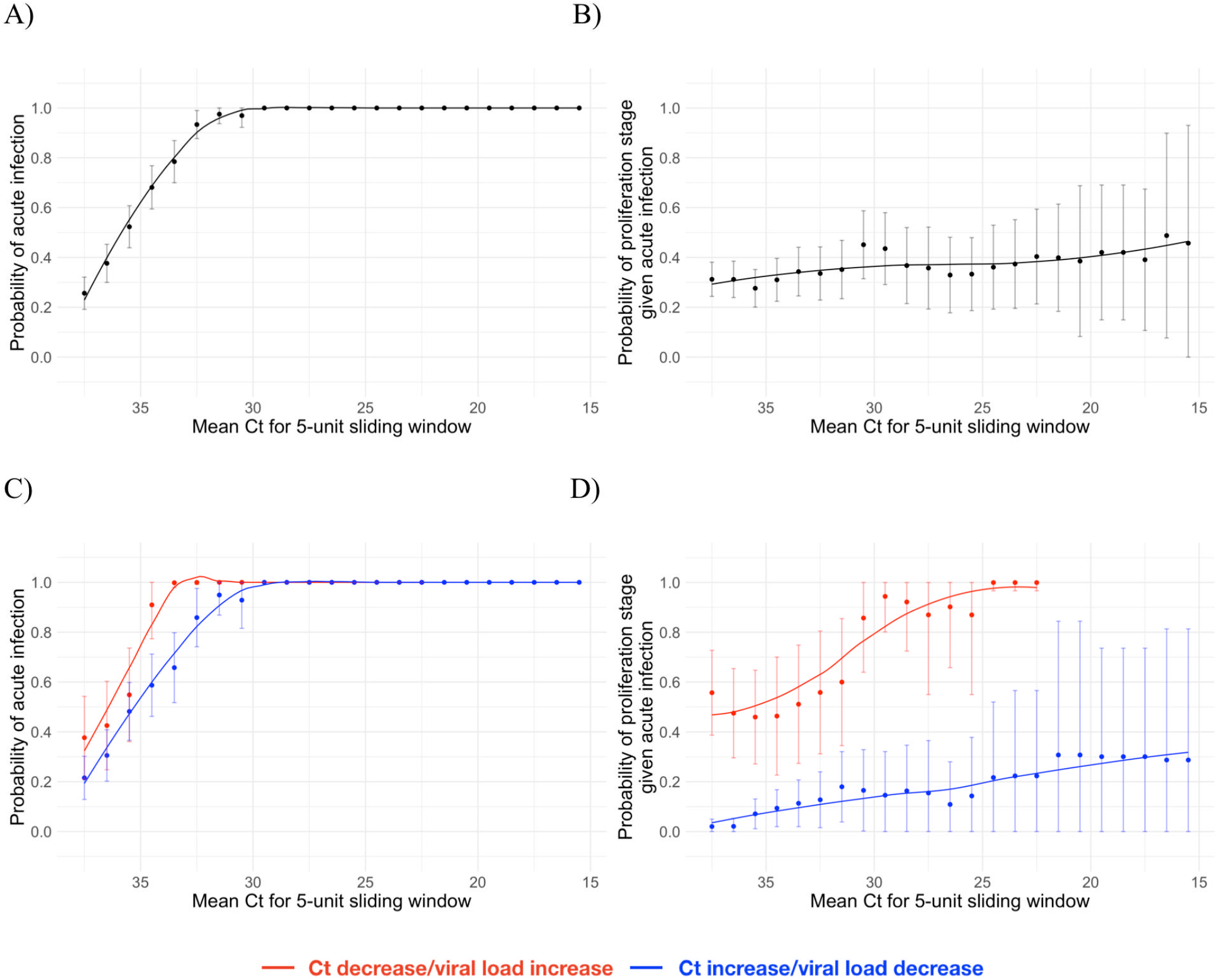
Relationship between single/paired Ct values and infection stage. Probability that a given Ct value lying within a 5-unit window (horizontal axis) corresponds to an acute infection (A, C) or to the proliferation phase of infection assuming an acute infection (B, D). Sub-figures A and B depict the predictive probabilities for a single Ct value, while sub-figures C and D depict the predictive probabilities for a positive test paired with a subsequent test with either lower (red) or higher (blue) Ct. The curves are LOESS smoothing curves to better visualize the trends. Error bars represent the 90% Wald confidence interval.

We assessed whether a second test within two days of the first could improve these predictions. A positive test followed by a second test with lower Ct (higher viral RNA concentration) was slightly more likely to be associated with an active infection than a positive test alone (**Figure 4C**). Similarly, a positive test followed by a second test with lower Ct (higher viral RNA concentration) was much more likely to be associated with the proliferation phase than with the clearance phase (**Figure 4D**).

We next estimated how the effective sensitivity of a pre-event screening test declines with increasing time to the event. For a test with limit of detection of 40 Ct and a 1% chance of sampling error, the effective sensitivity declines from 99% when the test coincides with the start of the event to 76% when the test is administered two days prior to the event (**Figure 5A**), assuming a threshold of infectiousness at 30 Ct^10^. This two-day-ahead sensitivity is slightly lower than the effective sensitivity of a test with a limit of detection at 35 Ct and a 5% sampling error administered one day before the event (82%), demonstrating that limitations in testing technology can be compensated for by reducing turnaround time. Using these effective sensitivities, we estimated the number of infectious individuals who would be expected to arrive at a gathering with 1,000 people given a pre-gathering screening test and a 2% prevalence of infectiousness in the population. Just as the effective sensitivity declines with time to the gathering, the predicted number of infectious individuals rises with time to the gathering (**Figure 5B**) since longer delays between the screening test and the gathering make it more likely that an individual will be undetectable at the time of testing but infectious at the time of the event. Changing the infectiousness threshold modulates the magnitude of the decline in effective sensitivity associated with longer testing delays; however, the overall trend is consistent (**Supplemental Figure 18**).

**Figure 5.**
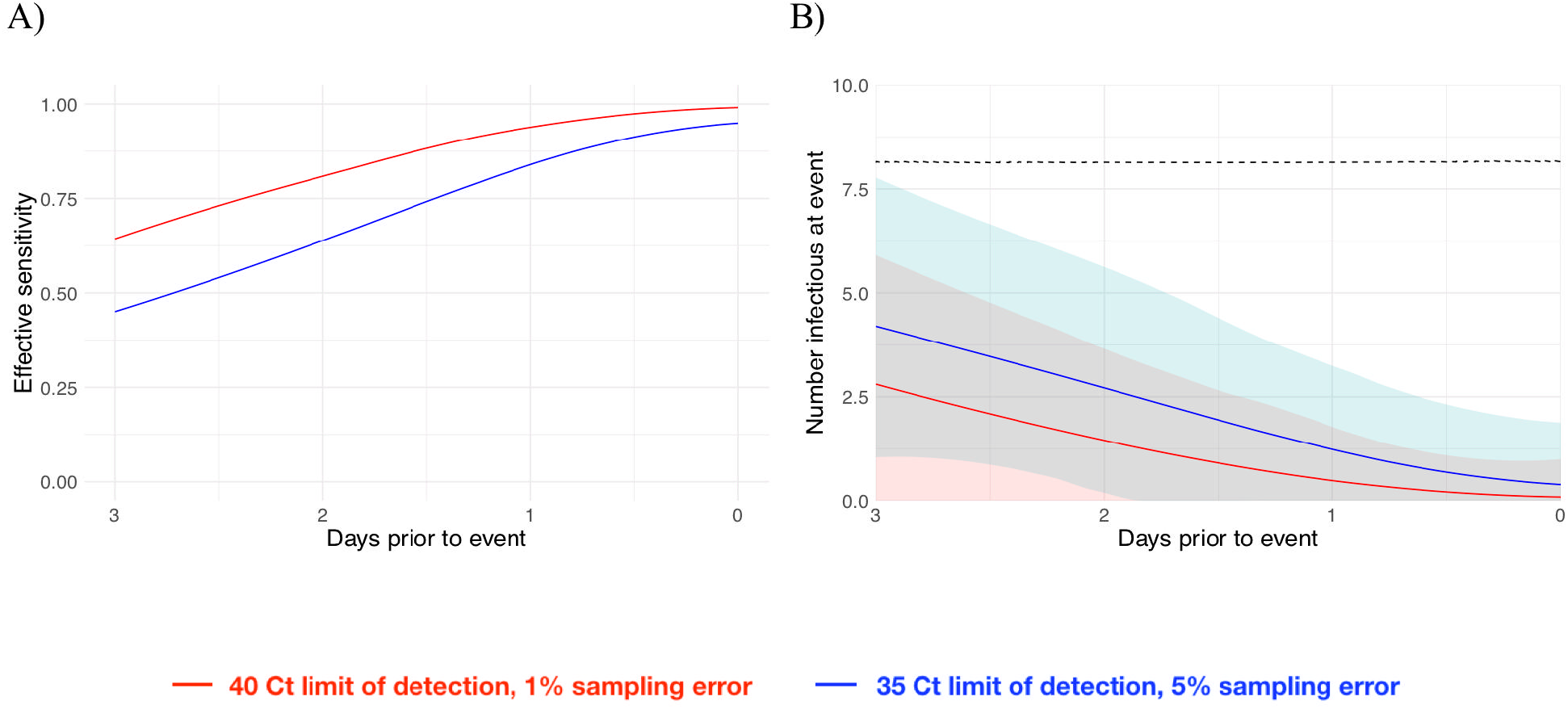
Effective sensitivity and expected number of infectious event attendees for tests with varying sensitivity. A: Effective sensitivity for a test with limit of detection of 40 Ct and 1% sampling error probability (red) and 35 Ct and 5% sampling error probability (blue). B: Number of infectious individuals expected to attend an event of size 1,000 assuming a population prevalence of 2% infectious individuals for a test with limit of detection of 40 Ct and 1% sampling error probability (red) and 35 Ct and 5% sampling error probability (blue). Shaded bands represent 90% prediction intervals generated from the quantiles of 1,000 simulated events and capture uncertainty both in the number of infectious individuals who would arrive at the event in the absence of testing and in the probability that the test successfully identifies infectious individuals. The dashed line depicts the expected number of infectious individuals who would attend the gathering in the absence of testing.

## Discussion

We provide the first comprehensive data on the early-infection RT-qPCR Ct dynamics associated with SARS-CoV-2 infection. We found that viral titers peak quickly, normally within 3 days of the first possible RT-qPCR detection, regardless of symptoms. Our findings highlight that repeated PCR tests can be used to infer the stage of a patient’s infection. While a single test can inform on whether a patient is in the acute or persistent viral RNA shedding stages, a subsequent test can help identify whether viral RNA concentrations are increasing or decreasing, thus informing clinical care. For example, a patient near the beginning of their infection may need to be isolated for different amounts of time than a patient near the end of their infection. If a patient is at risk for complications, closer monitoring and more proactive treatment may be preferred for patients near the start of infection than for those who are already nearing its resolution. We also show that the effective sensitivity of pre-event screening tests declines rapidly with test turnaround time due to the rapid progression from detectability to peak viral titers. Due to the transmission risk posed by large gatherings^11^, the trade-off between test speed and sensitivity must be weighed carefully. Our data offer the first direct measurements capable of informing such decisions.

Our findings on the duration of SARS-CoV-2 viral RNA shedding expand on and agree with previous studies^12–14^ and with observations that peak Ct does not differ substantially between symptomatic and asymptomatic individuals^3^. While previous studies have largely relied on serial sampling of admitted hospital patients, our study used prospective sampling of ambulatory infected individuals to characterize complete viral dynamics for the presymptomatic stage and for individuals who did not report symptoms. This allowed us to assess differences between the viral RNA proliferation and clearance stages for individuals with and without reported symptoms. The similarity in the early-infection viral RNA dynamics for both symptomatic and asymptomatic individuals underscores the need for SARS-CoV-2 screening regardless of symptoms. The progression from a negative test to a peak Ct value 2-4 days later aligns with modeling assumptions made in various studies^15,16^ to evaluate the potential effectiveness of frequent rapid testing programs, strengthening the empirical bases for their findings. Taken together, the dynamics of viral RNA shedding substantiate the need for frequent population-level SARS-CoV-2 screening and a greater availability of diagnostic tests.

Our findings are limited for several reasons. The sample size is small, especially with respect to symptomatic acutely infected individuals. The cohort does not constitute a representative sample from the population, as it was a predominantly male, healthy, young population inclusive of professional athletes. Viral trajectories may differ for individuals who have been vaccinated or who have been infected with SARS-CoV-2 variants, which we were unable to assess due to the timeframe of our study. Some of the trajectories were sparsely sampled, limiting the precision of our posterior estimates. Symptom reporting was imperfect, particularly after initial evaluation as follow-up during course of disease was not systematic for all individuals. As with all predictive tests, the probabilities that link Ct values with infection stages (**Figure 4**) pertain to the population from which they were calibrated and do not necessarily generalize to other populations for which the prevalence of infection and testing protocols may differ. Still, we anticipate that the central patterns will hold across populations: first, that low Cts (<30) strongly predict acute infection, and second, that a follow-up test collected within two days of an initial positive test can substantially help to discern whether a person is closer to the beginning or the end of their infection. Our study did not test for the presence of infectious virus, though previous studies have documented a close inverse correlation between Ct values and culturable virus^10^. Our assessment of pre-event testing assumed that individuals become infectious immediately upon passing a threshold and that this threshold is the same for the proliferation and for the clearance phase. In reality, the threshold for infectiousness is unlikely to be at a fixed viral concentration for all individuals and may be at a higher Ct/lower viral concentration during the proliferation stage than during the clearance stage. Further studies that measure culturable virus during the various stages of infection and that infer infectiousness based on contact tracing combined with prospective longitudinal testing will help to clarify the relationship between viral concentration and infectiousness.

To manage the spread of SARS-CoV-2, we must develop novel technologies and find new ways to extract more value from the tools that are already available. Our results suggest that integrating the quantitative viral RNA trajectory into algorithms for clinical management could offer benefits. The ability to chart a patient’s progress through their infection underpins our ability to provide appropriate clinical care and to institute effective measures to reduce the risk of onward transmission. Marginally more sophisticated diagnostic and screening algorithms may greatly enhance our ability to manage the spread of SARS-CoV-2 using tests that are already available.

## Data Availability

Code and data are available at https://github.com/gradlab/CtTrajectories

## Acknowledgements

We thank the NBA, National Basketball Players Association (NBPA), and all of the study participants who are committed to applying what they learned from sports towards enhancing public health. In particular, we thank D. Weiss of the NBA for his continuous support and leadership. We are appreciative of the discussions from the COVID-19 Sports and Society Working Group. We also thank D. Larremore for comments on the manuscript, J. Hay and R. Niehus for suggestions on the statistical approach and P. Jack and S. Taylor for laboratory support.

## Author contributions

SMK conceived of the study, conducted the statistical analysis, and wrote the manuscript. JRF conceived of the study, conducted the laboratory analysis, and wrote the manuscript. CM conceived of the study, collected the data, and wrote the manuscript. SWO conducted the statistical analysis. CT analyzed the data and edited the manuscript. KYS analyzed the data and edited the manuscript. CCK conducted the laboratory analysis and edited the manuscript. SJ conducted the laboratory analysis and edited the manuscript. IMO conducted the laboratory analysis. CBFV conducted the laboratory analysis. JW conducted laboratory analysis and edited the manuscript. JW conducted laboratory analysis and edited the manuscript. JD conceived of the study and edited the manuscript. DJA contributed to data analysis and edited the manuscript. JM contributed to data analysis and edited the manuscript. DDH conceived of the study and edited the manuscript. NDG conceived of the study, oversaw the study, and wrote the manuscript. YHG conceived of the study, oversaw the study, and wrote the manuscript.

## Competing interests

JW is an employee of Quest Diagnostics. JW is an employee of Bioreference Laboratories. NDG has a consulting agreement for Tempus and receives financial support from Tempus to develop SARS-CoV-2 diagnostic tests. SMK, SWO, and YHG have a consulting agreement with the NBA.

**Supplementary Information** is available for this paper.

